# Protocol for Compliance Assessment of the District Health Management Information System: Supervision of Data Management Processes for HIV Care in Public Healthcare Facilities in South Africa

**DOI:** 10.1101/2025.02.18.25322460

**Authors:** Eunice B. Turawa, Jeannine Uwimana Nicol, Duduzile Ndwandwe, Edward Nicol

## Abstract

As South Africa strives to meet the Joint United Nations Programme on HIV/AIDS (UNAIDS) 95-95-95, targets require adequate patient data to track people living with HIV (PLHIV) and link them to care. The National District Health Management Information System (DHMIS) Facility Report Standard Operating Procedures (SOP) aims to standardise data collection and use across health facilities. However, data quality issues persist after a decade of implementing the DHMIS-SOP. Currently, there is limited evidence on DHMIS-SOP compliance at the facility level. This study will explore healthcare managers’ compliance with DHMIS-SOP in HIV data management, identify facilitators and barriers to compliance, and develop strategies to improve adherence.

**Methods:** A multimethod study will be conducted in 46 selected HIV clinics in the uMgungundlovu District, KwaZulu-Natal. A scoping review will identify compliance frameworks for HIV data management. The PRISM Organisational Behavioural Assessment Tool (OBAT) and Management Assessment Tool (MAT) will assess 161 healthcare professionals’ knowledge of DHMIS-SOP content, motivations, and data supervision by the district management team. The results will inform in-depth interviews (IDIs) with 10-12 key informants to explore perceptions and experiences of DHMIS-SOP compliance and identify barriers and facilitators. A compliance framework will be developed to support SOP adherence in HIV data management.

**Analysis:** The scoping review will synthesise concepts and compliance frameworks/tools for SOP adherence. Descriptive statistics will report the percentages and proportions for categorical data, while mean, standard deviation (SD), median, and interquartile range (IQR) will report findings for continuous data. Frequency distributions, univariate and bivariate analyses, and multiple regression will describe and examine the strength of the relationship between facility managers and the district office on HIV data management processes, challenges and strategies for SOP compliance. For dichotomous data, 95% confidence limits will be calculated. A general inductive method driven by key barriers or facilitators of compliance will analyse the qualitative data. Two reviewers will independently translate transcripts and create a structured coding framework. The source quotes will be assessed and agreed upon, and a robust thematic report will be generated based on the combined analyses. A comparative analysis between and within interviews will develop coded information categories linked to broader themes evolving across interviews. This will guide data analysis in the ATLAS.ti software. Qualitative and quantitative data are integrated where appropriate. The findings of these phases will inform an expert panel workshop where an SOP compliance framework will be developed to improve SOP adherence in HIV data management in primary healthcare settings.

## Background

The Universal Test and Treat (UTT) strategy recommends that all at-risk populations screen for Human Immunodeficiency Virus (HIV) infection and individuals diagnosed as HIV positive receive early treatment irrespective of their CD4 count and clinical presentation ^**[1, 2]**^. The approach prioritises HIV testing, same-day antiretroviral therapy (ART) initiation (SDI), and prompt linkage-to-care (i.e., successful completion of a first clinic visit within two weeks of an HIV diagnosis, as documented in TIER.Net) ^**[3]**^. Even though ART treatment is freely available nationwide through the UTT strategy, challenges in linkage to HIV care persist, and a substantial proportion of patients are lost between the time of HIV diagnosis and the onset of HIV care and ART treatment ^**[4, 5]**^. Poor referral systems ^**[6]**^ and knowledge gaps in clinical guideline compliance ^**[7, 8]**^ fuel poor health services. Gaps in linkage to care and tracking of HIV-infected individuals who missed appointments or dropped out of care remain problematic^**[9]**^ because existing patient identification systems are paper-based, limiting patient tracking. Of concern is poor data quality (i.e., data that are not sufficiently accurate, timely, credible, and complete) because the information is poorly captured partly due to insufficient supervision and mentorship in HIV data management among healthcare facility managers ^**[10, 11]**^.

Compliance with standards in HIV data management processes and documentation is crucial to achieving the UNAIDS target and eradicating HIV/AIDS. Noncompliance with clinical data collection Standard Operating Procedures (SOP) leads to erroneous and misleading documentation in HIV services, fuelling low-quality services, billing discrepancies, and litigation ^**[12]**^, including morbidity and mortality rates ^**[13]**^.

Global medication error was estimated at $42 billion per year ^**[14]**^, while annual adverse events were close to 134 million, contributing to 2.6 million deaths in low and middle-income countries (LMICs) ^**[14]**^. The National Health Service (NHS) England pays close to £98.5 million annually for avoidable erroneous prescriptions in healthcare delivery ^**[15, 16]**^. The most significant errors include missed diagnoses, prescriptions, and low compliance with medical recording standards. In Indonesia, 60% of nurses evade SOPs in patient care ^**[17]**^, and Haug 2013 indicated that 67% of misdiagnoses and misinterpretations, erroneous prescriptions, and patient misidentifications are traceable to poor recording, which is often related to low compliance with approved data recording standards ^**[18]**^.

In Africa, data management inefficiencies are common, leading to disruptions in healthcare, particularly in public settings ^**[19–21]**^. For example, data breaches generate the highest per capita costs in South Africa’s health sector. An average data breach per capita cost of $1.16 and $3.06 million was reported for 2015 and 2019, respectively ^**[22]**^. These challenges are partly due to poor SOP compliance with established standards and guidelines for data management processes. The South African National Core Standards for Health Establishments assessment also revealed poor compliance with public service standards.

Generally, guidelines/SOPs/policies are hardly implemented as envisioned by policymakers ^**[23, 24]**^. Therefore, scholars proposed that simple and robust frameworks/tools be integrated into SOP to enhance compliance and achieve the policy/SOP goals ^**[25, 26]**^. A compliance framework will facilitate monitoring and minimise errors, providing motivations for SOP compliance and seamless organisation management ^**[27]**^.

SOPs are designed to provide clear and concise instructions on specific procedures. It controls and provides transparency and helps perform routine operations effectively. In addition to creating a culture of accountability by defining the roles and responsibilities of HCPs in healthcare service delivery, it serves as a reference point, minimising errors and ambiguities ^**[28, 29]**^. Noncompliance with the SOP is a risk factor for serious healthcare mismanagement and litigation. It leads to missed diagnoses and prescriptions and poor billing systems ^**[30]**^. Deviations from SOPs and disparities between SOP content and real-world implementation have been reported in developed and developing countries ^**[17, 21, 31, 32]**^. About 67% of clinical data challenges are traceable to human error and poor compliance with approved standards ^**[33]**^. The most significant errors include missed diagnoses, medication prescriptions, and low compliance with medical recording standards ^**[34]**^. In high-income countries, about one in ten patients experience harm during hospital care ^**[14]**^. Alhidayah (2020) suggests that only 14.7% of nurses comply with standard procedures for patient safety. In Africa, data management inefficiencies are common, leading to disruptions in healthcare, particularly in public healthcare settings ^**[19]**^.

The reasons for deviation range from a lack of resources, organisational and technical challenges, SOP inadequacy or not being user-friendly, low user capacity, complexities of applicable technologies, non-standardised data coding system, lapses in managerial oversight, and delayed decision-making to poor SOP compliance ^**[32, 35–40]**^.

Generally, there is a decline in HIV epidemics worldwide, but significant work predicated on accurate and reliable data is still needed, especially among the young population. South Africa bears the highest HIV burden in the World, with approximately 7.7 million people living with HIV(PLHIV), culminating in a national prevalence of 12.7%, 16% in KwaZulu-Natal province, and 19.5% in uMgungundlovu district in 2022 ^**[41–43]**^.

Although ART is widely accessible nationwide through the UTT policy, the effectiveness of the policy is often compromised by patient attrition between HIV diagnosis and antiretroviral treatment (ART) initiation^**[4, 5]**^. The reasons for these gaps include incomplete and inconsistent patient data records, poor referral systems, and knowledge gaps in clinical guidelines and compliance. Of concern are poorly captured and incomplete patient data and insufficient supervision of HIV data management processes among health facility managers ^**[11]**^, which results in poor compliance in data recording and misinterpretation of information on HIV status ^**[44] [45]**^.

Frameworks are structured tools designed to help ensure compliance with organisational regulations and SOPs. They enhance adherence to SOPs and improve medical documentation and record-keeping. By clearly defining roles and responsibilities for compliance and integrating compliance controls into management processes, SOP frameworks contribute to overall compliance, ultimately improving health outcomes ^**[46–48]**^.

The South African District Health Information System is a “system that derives a combination of health statistics mainly from RHIS used in the public sector to monitor health services”^**[49]**^. The DHMIS policy (2011) aims to guide public health information management in health facilities in the country, strengthen health system monitoring, and enhance information use in healthcare planning by standardising data collection, dissemination, and use ^**[50]**^. It is an overarching national policy with associated processes, SOP, and data management standards in public health facilities.”TIER.Net (national HIV/ART electronic database) is embedded in the DHIS2 software and collects patient-level information at the health facility level to monitor epidemic trends and provide information for timely linkage of PLHIV to care and treatment ^**[51]**^. The DHMIS-SOP was enacted in 2012/13 and updated in 2016 for all public health facilities and districts to guide data management nationwide ^**[28, 52]**^. The SOP (DHMIS-SOP, 2016) aim to strengthen DHIS performance ^**[28]**^ by focusing on common data quality challenges and procedures, including HIV data processing. The DHMIS-SOP 2016 outlined the roles and responsibilities of HCWs in SOP use for standardising data management processes. However, after a decade of implementing the DHMIS-SOP, persistent data challenges and gaps remain a cause for concern in health services. Although HIV data processing and quality have significantly improved over the years, gaps in quality and capturing errors remain fundamental limitations, especially at the facility level ^**[53, 54]**^. The literature suggests that DHMIS-SOP was implemented in only 65% of public healthcare facilities, and only 55% of HCWs were trained in SOP use ^**[54]**^. Murphy 2013 highlighted the absence of DHMIS-SOP documents and the non-implementation in HIV data management ^**[55]**^. Although the Western Cape Province has the most complete clinical data, within this seemingly complete dataset, notable duplicate records, accounting for 10% to 20% of entries, are reported ^**[56]**^. The degree to which HCWs comply with the DHMIS-SOP in HIV data management processes is unknown ^**[57]**^. Enormous resources and funding have been dedicated to HIV programmes in South Africa. These initiatives have improved the TIER.Net performance and data quality compared to other in-facility data sources. Yet, the system is not totally free of inconsistency, misclassifications, misreported data, and missing outcomes ^**[53]**^.

Generally, policies, guidelines, and SOPs are hardly implemented as envisioned by policymakers ^**[23, 24]**^. Therefore, scholars have proposed simple, robust frameworks/tools to enhance implementation and compliance to achieve the policy/SOP goals ^**[25, 26]**^. Integrating a compliance framework into an SOP facilitates compliance and effective monitoring, minimises errors, and aids seamless organisation management ^**[58–60]**^.

Fig 1 illustrates a causal loop diagram (CLD) with the interconnected relationships and feedback loops between system variables in compliance. It illustrates how variables influence one another, with arrows indicating the direction of impact. The circular arrows depict reinforcing (R) loops, which promote positive behaviours, and balancing loops, which can stabilise or counteract behaviours related to DHMIS-SOP compliance in HIV data management processes.

**Fig 1.**
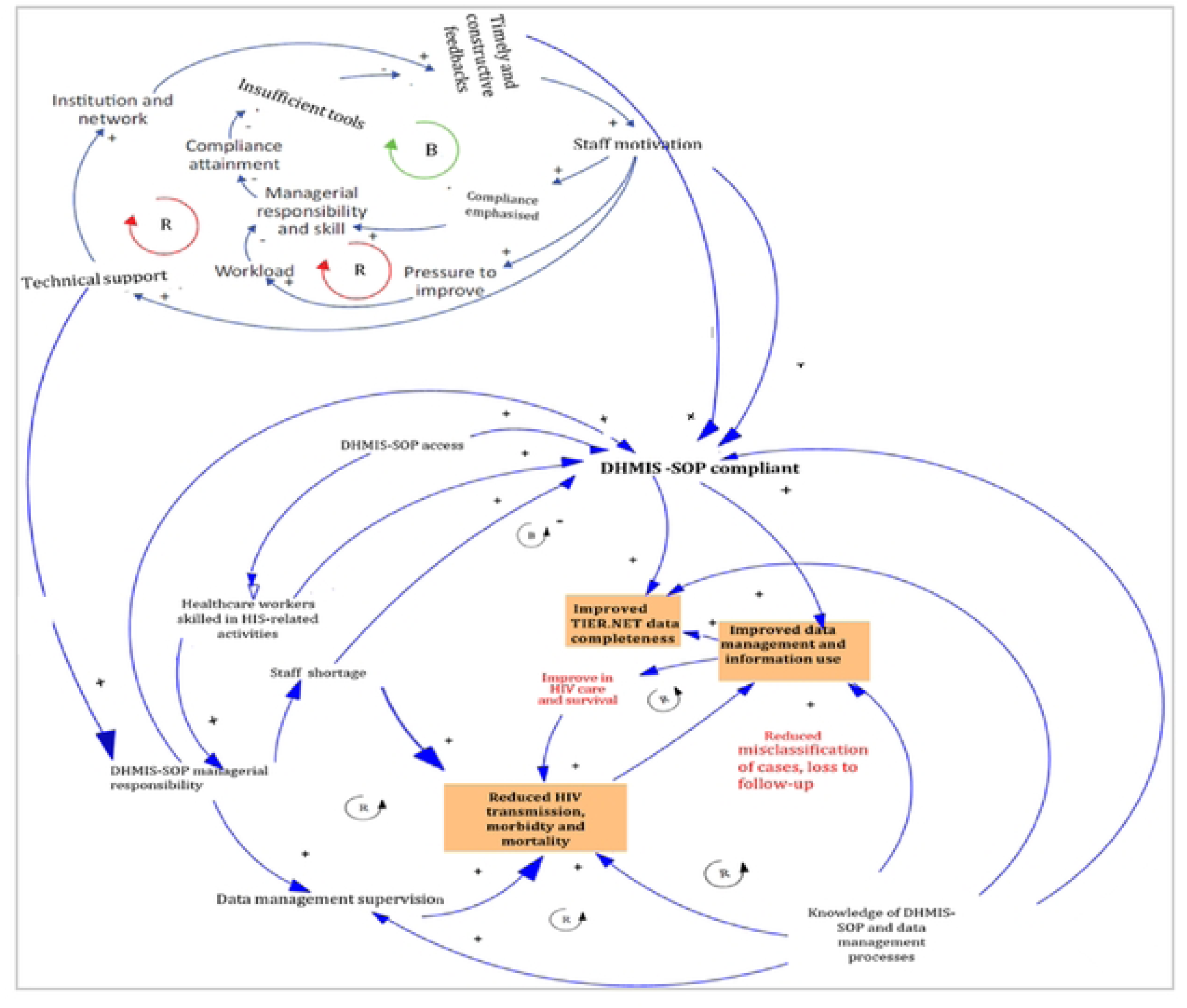
Critical factors influencing DHIS performance and routine HIV data management at the facility level.

The DHMIS-SOP was enacted to strengthen data management and information use. Currently, there are concerns about poor DHMIS-SOP compliance in many facilities, with reports of poor implementation and compliance across health facilities ^**[10, 54, 55, 61]**^.

The evidence of DHMIS-SOP effectiveness in improving RHIS quality is unclear, and it is difficult to ascertain whether the SOP is achieving its goals. The information, communication, and technology (ICT), data quality assurance tools within DHIS2 and integrated TB/HIV data management SOP ^**[62, 63]**^ have not yet yielded the expected preparedness in HIV data management processes. Few studies have referenced DHMIS-SOP on inconsistency data ^**[54, 61, 64]**^. Asah (2021) compared leadership styles in the Community of Information Practice (COP) ^**[64]**^; the MEval-SIFSA project reported shortfalls in managers’ HIS skills and information use per DHMIS-SOP ^**[54]**^. On the other hand, Nicol (2015) identified gaps in SOP and proposed further research on organisational factors that affect the performance of RHIS^**[61]**^. None of these studies examines the roles and responsibilities of facility managers in steering DHMIS-SOP compliance at the facility level for better data quality. The HIV data management challenges, despite the considerable investment in RHIS and HIV services, raise fundamental questions such as: Why has DHMIS-SOP not succeeded in standardising facility data? Are clinical staff adequately skilled and trained to use DHMIS-SOP? Is DHMIS-SOP user-friendly or overly complex? Are there enough resources to implement SOP? What are we not doing right? These are some of the burning questions the current study will investigate.

Thus, the proposed study will evaluate DHMIS-SOP compliance using the HIV programme as a case study and evaluate the extent to which the facility managers perform their roles and responsibilities in HIV data management processes. It will identify facilitators of /barriers to DHMIS-SOP compliance, provide valuable information on the challenges and gaps in SOP compliance at different health system levels, and provide strategies to improve compliance, thereby supporting the National Development Plan (NDP) goal of ending HIV/AIDS epidemic by 2030. Assessing the TIER.Net will reveal how well or poorly other in-facility data sources perform.

## Materials and methods

### The main objectives of the study include

1. To identify and describe compliance tools and frameworks for HIV data management processes.
2. To describe the extent to which healthcare managers in the uMgungundlovu district comply with the DHMIS-SOP for the HIV data management processes.
3. To explore facility managers’ views and perceptions on compliance with the DHMIS-SOP for HIV data management processes.
4. Develop a compliance framework and tools to assist managers to comply with the HIV data management processes at the district and facility levels.

### Study Setting

The study setting for this study has been described extensively in another peer-reviewed publication ^**[65]**^. The study will be conducted in the rural uMgungundlovu district of KwaZulu-Natal (KZN) province of South Africa. The district comprises seven sub-district strata): Msunduzi, uMshwathi, uMngeni, Richmond, Mkhambathini, Mpofana, and Impendle. The health district has one state-aided clinic, 17 mobile clinics, and 46 Primary Health Care (PHC) facilities staffed with HCWs to provide HIV services. This district is one of the National Health Insurance (NHI) pilot sites, with about 84% of the population being medically uninsured. ^**[66]**^ Inadequate HIV data management processes and poor paper-based filing systems characterise the district.^**[67]**^ The RITSHIDZE report highlights the need to strengthen data processing for tracing and tracking adolescent girls, young women, and men (AGYW&M) in HIV services within the district.^**[67]**^ These factors and reports make uMgungundlovu an ideal setting for assessing DHMIS-SOP compliance in HIV data management processes and quality performance.

## Methodology

### Study design

A multimethod design ^**[68]**^ will investigate how facility managers steer SOP compliance at primary health facilities in the uMgungundlovu district. This design will provide cross-validation and corroboration, adding credibility to the study findings ^**[69]**^. A scoping review will be conducted to identify and describe compliance tools and frameworks related to HIV data management processes (**Objective 1**). A cross-sectional survey will examine the roles of facility managers in promoting DHMIS-SOP compliance for good quality HIV data management (**Objective 2**). The results of this phase will inform data collection for the qualitative phase, where In-depth interviews (IDI) will be conducted to gain insights into the perceptions, views, and experiences of facility managers regarding DHMIS-SOP compliance. These interviews will explore the barriers to and facilitators of compliance, as well as strategies to mitigate these barriers. (**Objective 3**). The study findings will be integrated during the result interpretation phase. Subsequently, an expert workshop will identify and map compliance concepts and develop a compliance framework to enhance HIV data management (**Objective 4**). Table 1 provides an overview of study designs and data collection methods.

**Table 1.** The study methods and data collection tools for SOP compliance study.

|  | Objective 1 | Objective 2 | Objective 3 | Objective 4 |
| --- | --- | --- | --- | --- |
| **Data collection tool** | To identify and describe compliance tools and frameworks for HIV data management processes. | To describe the extent to which healthcare managers comply with the DHMIS-SOP for HIV data management processes. | To explore facility managers' views and perceptions on DHMIS-SOP compliance for HIV data management processes. | To develop a compliance framework and tools to assist managers comply with the HIV data management processes. |
| **Literature review and analysis of secondary data** |  |  |  |  |
| **Scoping Review:**<br>Secondary data analysis | XX |  |  |  |
| **Quantitative methods** |  |  |  |  |
| **Cross-sectional study:**<br>*The PRISM's performance diagnostic tool (Use of information module, OBAT, and MAT).* |  | XX |  |  |
| **Qualitative methods** |  |  |  |  |
| **In-depth interviews:**<br>*USAID policy-implementation tool.* |  |  | XX |  |
| **Expert group workshop** |  |  |  | XX |

## Study Population and Recruitment

The study population and recruitment will be based on the various study components to answer the research questions adequately as described in Table 2.

**Table 2.** Study population and sample size from fixed health facilities in uMgungundlovu Health District, Kwa-Zulu-Natal Province.

| Categories of healthcare providers | Facility Level | Sub-district Level | District Level | Number to be interviewed |
| --- | --- | --- | --- | --- |
| Facility managers | 46 | - | - | 46 |
| Professional Nurse | 46 | - | - | 46 |
| Facility information officer | 46 | - | - | 46 |
| HAST Coordinator | - | 7 | - | 7 |
| PHC Manager | - | 7 | - | 7 |
| Sub-district manager | - | 7 | - | 7 |
| District information officer | - | - | 1 | 1 |
| District manager | - | - | 1 | 1 |
| **Total** | **138** | **21** | **2** | **161** |

\*Healthcare provider categories and list extracted from "Strengthening Health System's Capacity for Linkage to Care (SHeS'Cap) for AGYW and men in South Africa" Project. A Continuation of Funding Opportunity Announcement CDC-RFA-GH13-1340, "Strengthening Strategic Information, program implementation and Capacity Building to reduce morbidity and mortality in South Africa under the President's Emergency Plan for AIDS Relief (PEPFAR)" Project.

**Table 3.** Eligibility criteria for article inclusion in the scoping review.

| Inclusion criteria | Exclusion criteria |
| --- | --- |
| Studies published in peer-reviewed journals describing SOP compliance tools or frameworks for HIV data management processes and analysis will be eligible for inclusion.<br>Data describing tools for HIS-data management tasks will be eligible for inclusion. | Studies that do not focus on compliance frameworks for HIV data management will be excluded. |
| Studies published in English and results are available for synthesis. | Studies, not English Language<br>Abstracts with empirical data/results that cannot be synthesized. |
| Studies published from January 2000 to the present. | Studies published before January 2000. |

The scoping review will draw on publicly available datasets to identify and describe available SOP compliance frameworks that can be adapted to improve DHMIS-SOP compliance in HIV data management processes. In contrast, the target population for the quantitative and qualitative components of the study will come from purposively selected 46 fixed health facilities, all seven sub-districts, and the health district office. The study participants will include HCWs with existing roles of Facility managers, HAST coordinators, PHC managers, Clinical nursing practitioners (CNP), Facility health information officers/data managers, the district manager and sub-district managers listed in Table 2. These individuals are involved in HIV data management processes and supervision according to DHMIS-SOP; therefore, they are best positioned to provide detailed reports on DHMIS-SOP compliance and associated challenges.

### Expert group

A team of experts in health information systems, HIV data management, and facility managers will be selected. They will receive invitations to a workshop on developing a DHMIS-SOP compliance framework for managing HIV data processes at the facility level.

## Sampling

An overview of the research components is illustrated in Fig 2.

**Fig 2.**
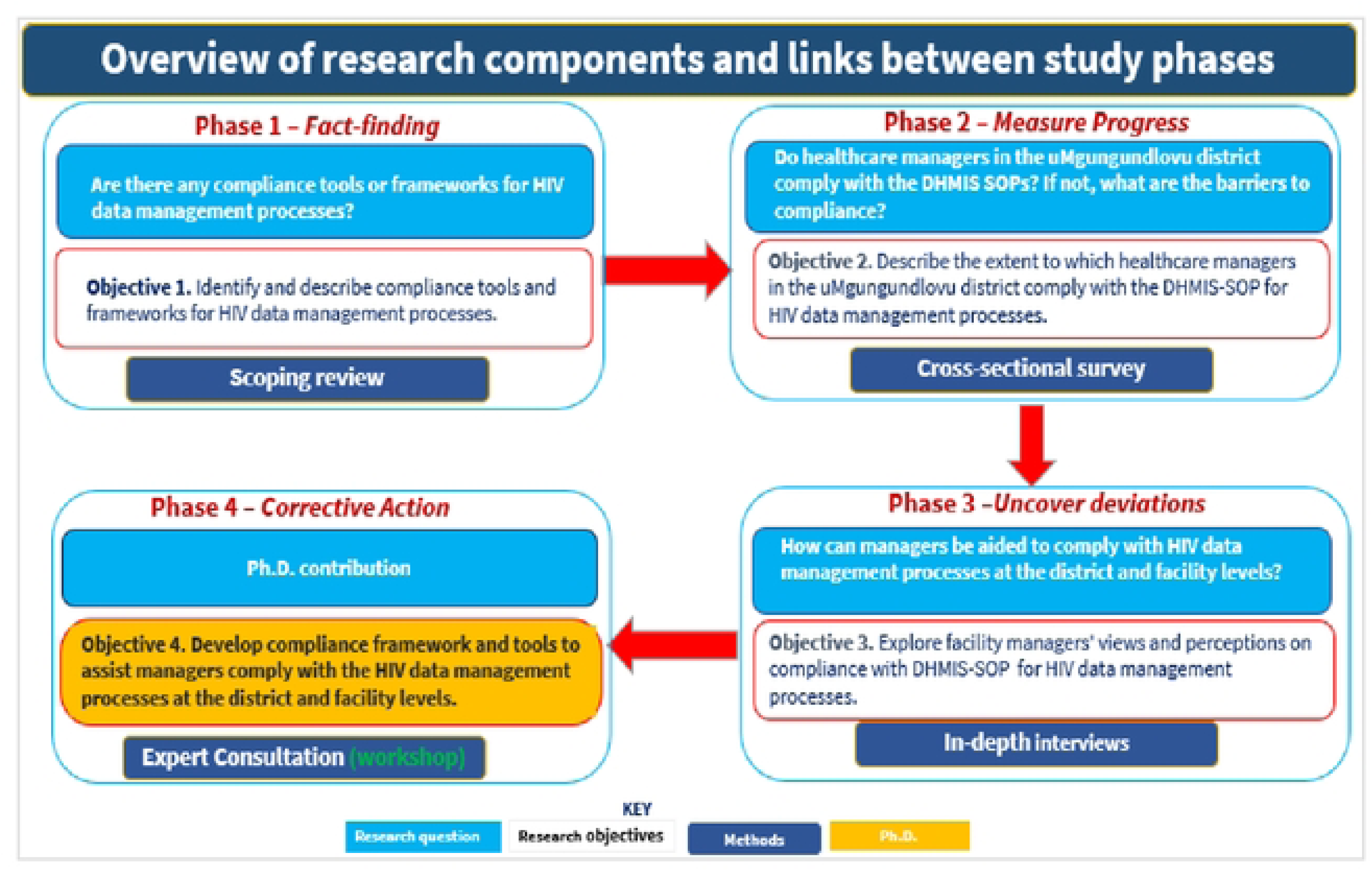
Overview of research components, data collection methods, and links between study phases.

**<u>The scoping review</u>** will follow Arksey and O’Malley’s framework ^**[70]**^ and the JBI Manual for Evidence Synthesis scoping review process^**[71]**^. A standard protocol will guide the review process, and a systematic search of scientific databases such as PubMed, Scopus, and Web of Science will be performed to identify articles on compliance frameworks/tools articles. Articles retrieved from databases will be exported into EndNote 20 software, where articles will be deduplicated and exported into an Excel spreadsheet for further independent screening of titles and abstracts by two review authors (Eunice Turawa [ET] and Research Assistant [RA]) for eligibility for inclusion. Potential eligible articles will be assessed for quality, scientific rigour and completeness of data. The Critical Appraisal Skills Programme (CASP) qualitative checklist will evaluate each article for scientific soundness, especially for qualitative studies.^**[72]**^ A PRISMA flow diagram will present the article search and selection processes. A “Table of Characteristics of Included Studies” will also provide detailed information about the study methods and quality of articles included in the study. Discrepancies between researchers will be resolved through discussion or intervention by the study supervisors. The number of excluded articles at each selection stage will be noted, including reasons for exclusion.

**<u>Cross-sectional survey</u>** (*Objective 3*). A purposive sampling method will enrol 161 HCWs (at least one per facility) in existing roles as facility managers, HAST coordinators, PHC managers, Clinical Nursing Practitioners (CNP), and facility health information officers/data managers from all 46 fixed health facilities, as shown in Table 2. The district manager and all sub-district managers from all seven local municipalities, including HCWs involved with data management activities at the health facilities, who agree to participate in the study will be sampled.

**<u>In-depth interview</u>** (*Objective 3*). This will comprise 10-12 purposively selected key informants from the list of HCWs listed in Table 2. They will provide information on their views and perceptions of DHMIS-SOP compliance in HIV data management via an in-depth interview. Participants will provide information on barriers and facilitators of DHMIS-SOP compliance, strategies to overcome obstacles, and practical approaches to improve compliance in HIV data management.

For cross-sectional study and the in-depth interview, all necessary arrangements regarding the date and time of the interview and venue will be secured. Before the interview, the researcher will visit the health facilities and participants to explain the importance of the interview and the process. Participants will also consent to audio recording and will keep their responses confidential.

**<u>Expert group</u>** (*Objective 4*). A team of experts in health information systems, HIV data management, and facility managers will be selected. They will receive invitations to a workshop on developing a DHMIS-SOP compliance framework for managing HIV data processes at the facility level.

Using participatory purposive sampling, 10-12 experts will be sampled for this study component. This will include at least one representative of the National Health Information System of South Africa (NHISSA) from each Province, a member of the uMgungundlovu Health District Executive Committee, local health information system stakeholders, facility managers, and health information managers. The expert panel will leverage the findings from the previous phases of the study and examine the contents of the DHMIS-SOP to guide the development of a compliance framework which can improve DHMIS-SOP adherence in HIV data management processes. Fig 2 provides an overview and links between study phases.

## Data collection and analysis

### Scoping review

Two reviewers [ET and RA] will extract relevant demographic data (name of the first author and year of publication, study titles, country, study setting and population, study objectives, and study citation for each article). Study methods, type of tool/framework, method of assessment, and outcome assessed, including comments and valuable notes, will be extracted for clarity. Data will be exported into an Excel spreadsheet for synthesis. The outcomes will consolidate into specified theme and sub-theme headings to produce a refined list of themes and sub-themes while retaining essential information from each article. Data will be summarised according to key compliance concepts and frameworks/tools, including the context in which the tools were used, what outcomes were measured, and key findings. A descriptive narrative synthesis and tabulations will summarise key themes/sub-themes, patterns, or improvements in SOP compliance. The relationship between health facility type (hospitals, PHC, day clinics) and impacts on compliance will be noted. To minimise potential biases in data interpretation, at least three experienced researchers will examine the themes and sub-themes, including merging codes. This will provide an overview of available frameworks/tools that can be adapted for SOP compliance in HIV data management. Throughout the study processes, discrepancies between reviewers will be resolved through discussion and the involvement of a third reviewer.

***For the cross-sectional study***, pretested MAT and PRISM Diagnostic tools ^**[73]**^ will elicit data on the supervision of HIV data management processes per DHMIS-SOP, staff training processes, availability of SOP, and open display of SOP to guide data management in the facility collected. The OBAT ^**[73]**^ will examine and provide data on managerial knowledge, skills, and staff motivation and feedback. The frequency and timeliness of supervision and feedback between the sub-district Data Management Team (DMT) and the facility and facility managers to facility staff will be investigated. Strategies and initiatives taken to improve HIV data management processes will be collected.

Data analysis will include quantitative descriptive statistics such as frequency distribution describing participants’ background characteristics. A bivariate analysis will be conducted, and a cross-tabulation of participants’ characteristics with behavioural and organisational factors (such as managerial roles, leadership, and supervision of HIV data collection per DHMIS-SOP) will be performed to explore the significant effect on SOP compliance in HIV data management processes.

Results will be displayed using bar charts, tables and figures. Pearson’s chi-square test will detect association. The pairwise correlation test will assess the relationship and impact of SOP compliance on HIV data management, including significance levels (95% confidence level) and association in the analyses. All analyses will be performed using STATA version ^18^ ^**[74]**^.

### In-depth Interviews

A pretested tool will explore and elicit responses from participants regarding their views and perceptions of DHMIS-SOP compliance for HIV data management processes. A pretested interview guide will ensure adequate information is collected, with interviews lasting 30 to 45 minutes. Kvale (1994) ^**[75]**^ recommended steps for IDI will be followed during data collection. The method offers an opportunity to capture rich, descriptive data about managerial attitudes, views, and lived experiences on SOP compliance, including a deeper understanding of the reasons for noncompliance and strategies for improving SOP adherence.

Audio recordings will be transcribed verbatim and analysed using thematic analysis. ET and RA will independently read each translated transcript and generate a structured coding framework based on the study objectives. Initial coding will be done descriptively, using a general inductive method driven by central research questions, focusing on significant themes to guide data analysis.^**[76]**^ Data will be analysed for consensus, disagreement, or divergences. Emerging themes, categories, and patterns will be noted.

A second-stage analysis will compare findings across the dataset, refining themes and developing higher-level categories. A comparative analysis between and within interviews assessment will be conducted, and sets of coded information linked to broader themes evolving across interviews will be generated.^**[76]**^ These codes will be used to analyse the data within the ATLAS.ti software, reflecting respondents’ views and perceptions, barriers, and strategies for improving SOP compliance in HIV data management.

The research team will triangulate the themes, and the final themes and sub-themes will be generated through group consensus and documented according to the consolidated criteria for reporting qualitative studies (COREQ) ^**[77]**^. Data triangulation will be performed in the study report section when possible.

## Developing compliance framework for SOP compliance

A one-day workshop via Zoom or face-to-face meeting will be organised to develop a consensus-based compliance framework for DHMIS-SOP compliance, as illustrated in Fig 3. Participants will review findings from the different components of the study (Phase 1, 2, and 3). An excerpt detailing facility managers’ roles and responsibilities per DHMIS-SOP will be shared with them. Additional components of the health information system and ICT essential for HIV data management at the facility level will be provided to ensure the developed framework suits facility settings. Experts will have four weeks to review these documents, identify keywords, and map concepts to develop a DHMIS-SOP compliance framework for improving HIV data management processes at the healthcare facilities in the uMgungundlovu district.

**Fig 3.**
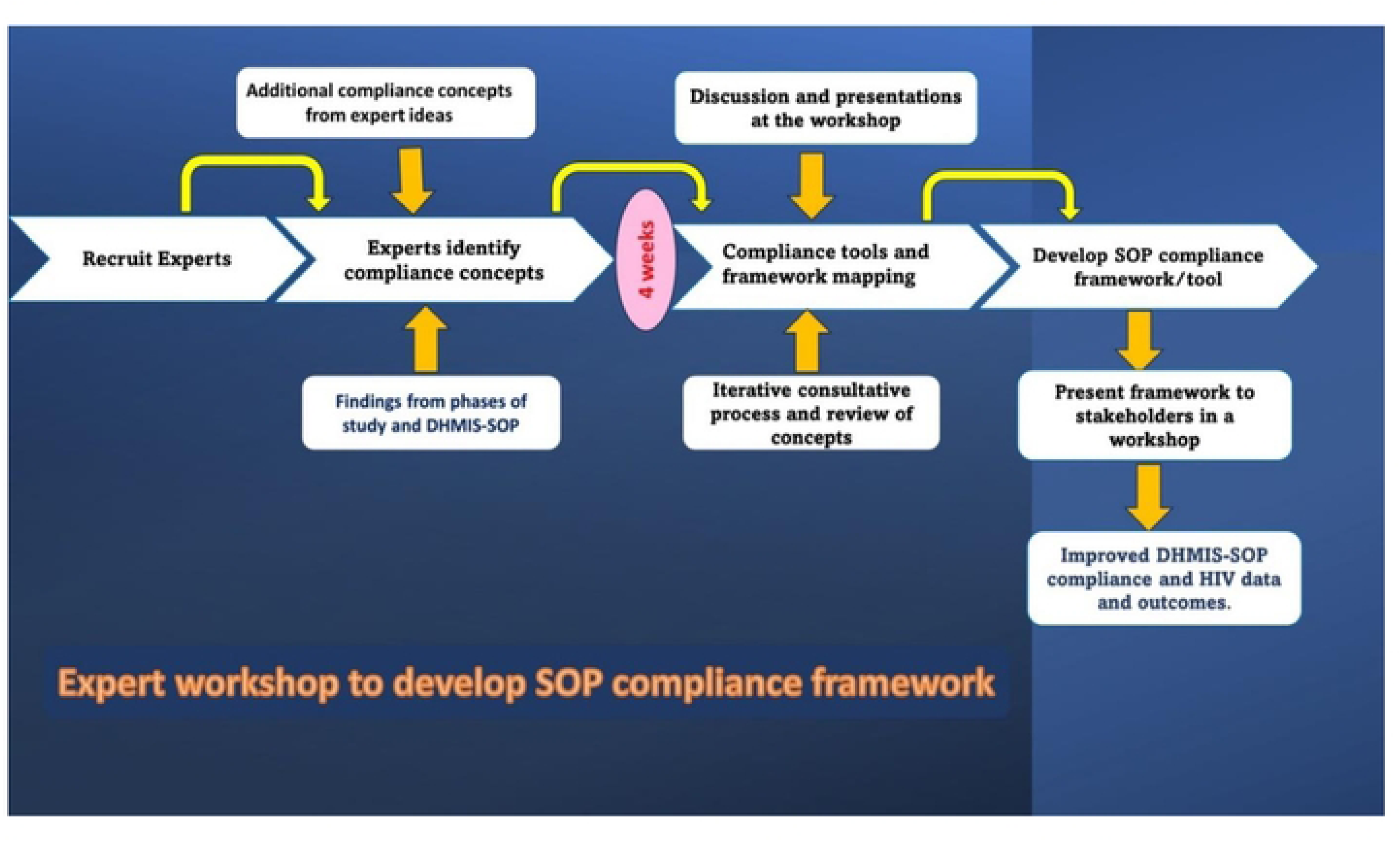
Expert consultation process for DHMIS-SOP compliance framework for HIV data management processes in uMgungundlovu, KZN, South Africa.

An open interactive group discussion will identify and collate concepts relevant to the healthcare facilities in the uMgungundlovu district. This will inform the first draft of the SOP framework. Experts can remove or add more concepts to the draft appropriately. In addition, identified concepts will be aligned with the prescribed roles and responsibilities of HCPs and facility managers as detailed in the DHMIS-SOP. Suggestions on appropriate concepts and context-suitable compliance framework generated from the workshop will be reviewed and applied to the drafted DHMIS-SOP framework. This process will ensure clarity, scientific rationale, and the framework’s validity.

### External review of developed framework

The framework will undergo peer review by two external reviewers who are experts in SOP framework development and implementation. The NHISSA committee members will appoint potential external experts and copies of the framework, and a clear guideline on the review process, criteria, and expectations will be shared with them. The exercise will broaden the perspective and validity of the developed tool. Feedback from external consultations will be carefully reviewed by the team and incorporated into the draft by consensus. The final version of the framework will be approved by consensus and presented to the provincial authorities, district, sub-district, and facility managers for endorsement.

## Data management

REDCap database software will build and manage data collection processes for the Project’s quantitative and qualitative components. The EndNote 20 database will manage database search output and initial study screening for the scoping review. We will follow the PRISMA-ScR checklist^**[78]**^ to process and synthesise the frameworks and/or tools.

## Ethics and dissemination

The Health Research Ethics Committee of Stellenbosch University has granted ethics approval (HREC/UREC reference number: S24/07/176) for the study to be conducted.

### Informed consent

Before data collection in study phases involving humans, consent forms will be applicable, and the study will be conducted according to ethics guidelines. We will obtain oral and written informed consent from all potential participants before they participate in the study. Participation is entirely voluntary, and during recruitment, participants will be informed that they can withdraw at any time without any negative consequences from the study team or the facilities involved. Any serious adverse event (e.g., breach of participant confidentiality) will be systematically documented and handled according to the Standard Operating Procedures of the Stellenbosch University Ethics Committee. This includes reporting adverse or unexpected events to the Committee within 48 hours. Participation in the study may involve disclosing emotional and personal matters, such as inadequate supervision, delayed reports, and feedback between HCWs or facility levels.

## Dissemination of study findings

The study results will be widely disseminated through scientific channels (such as publications and conference presentations), targeted feedback, and materials for the KZN Provincial Department of Health, the health facilities, district, and national levels. The study findings will be shared with key stakeholders, including those at the facility-based, district, provincial, and national levels. Outputs will be communicated through publications in scientific journals (e.g., BMJ Open), seminars, conferences, and Stellenbosch University Academic Day. The anonymity and confidentiality of the participants will be preserved by not revealing any identifying information while disseminating study findings. Outputs will include a recommendation for the DHMIS-SOP update based on summary findings and peer-reviewed journal articles.

### Study rigour

The quantitative study focuses on validity and reliability, while the qualitative component maintains credibility, dependability, confirmability, and transferability^**[79]**^. Close observation and triangulation will be performed, and multiple data collection methods will be used for the study. A range of informants and data verification processes will be in place, including an audit trail of all data, methodologies, and decisions at each research stage. The researcher will self-reflect to avoid personal bias and ensure the study results emerge from the collected data, not her predispositions. Dependability will be preserved by reporting the study processes and findings according to the COREQ checklist ^**[77]**^.

### Validity and reliability

A data management plan will be developed for each study phase to ensure good data quality and reproducibility. Patterns of missingness will be statistically assessed, noted, and reported. Data will be compared with published literature to ensure that self-reported data are meaningful. Random measurement errors are less likely as the questionnaires were validated.

### Protocol peer review

The current protocol was submitted to the SAMRC Scientific Committee and presented to the Stellenbosch University PhD committee for scientific review. Both Committees are satisfied with the protocol’s scientific merit.

## Study limitations and strengths

This study is the first to systematically evaluate DHMIS-SOP compliance, focusing on leadership, management, and supervision. The Project will identify barriers/facilitators of SOP compliance and develop a compliance framework to help facility managers/leaders steer SOP compliance in data collection. Each study objective will be systematically investigated, and a data management plan will be formulated to ensure the data quality and validity of the study findings. The key limitation of this research is that the results will not be generalisable to other provinces in South Africa, as SOP compliance may differ significantly.

## Discussion

This study aims to provide evidence to guide efforts to improve DHMIS-SOP compliance in HIV data management by analysing how facility managers have facilitated DHMIS-SOP in HIV data management processes. It also seeks to identify the barriers to and factors that encourage compliance and develop an SOP compliance framework to enhance the utilisation of DHMIS-SOP.

The study is crucial in supporting South Africa’s efforts to meet the UNAIDS 95-95-95 targets for HIV care. Achieving these goals requires effective HIV data management to trace and track PLHIV, ensuring they are linked to the appropriate care and services. The National District Health Management Information System (DHMIS) Facility Report SOP are designed to standardise data collection and usage across health facilities. This study will fill a critical gap by evaluating the extent to which healthcare managers at the facility level comply and steer DHMIS-SOP compliance in HIV data management to improve HIV data completeness and consistency for effective monitoring and tracking of PLHIV, allocate resources efficiently, plan and evaluate the impact of HIV programmes in the uMgungundlovu district.

The findings will contribute to understanding how DHMIS-SOP compliance can be enhanced, its importance in improving HIV data management processes, and ultimately achieving the UNAIDS targets. Developing a compliance framework will guide facility managers in improving SOP adherence, ensuring that data management processes at the facility level support better outcomes in HIV care and treatment. The importance of this study lies in its potential to strengthen the foundational data systems crucial for achieving national HIV health goals and improving the overall effectiveness of HIV management in South Africa.

## Data Availability

None

## Acknowledgements

Not applicable

## Contributorship

- Eunice Turawa (ET) and Edward Nicol (EN): Conceptualization, Methodology, and Writing the original draft.
- Jeannine Uwimana Nicol (JUN) and Duduzile Ndwandwe (DN): Review & editing, contribute to the methodology.

